# Multiple Introductions of SARS-CoV-2 Alpha and Delta Variants into White-Tailed Deer in Pennsylvania

**DOI:** 10.1101/2022.02.17.22270679

**Authors:** Andrew D. Marques, Scott Sherrill-Mix, John K. Everett, Hriju Adhikari, Shantan Reddy, Julie C. Ellis, Haley Zeliff, Sabrina S. Greening, Carolyn C. Cannuscio, Katherine M. Strelau, Ronald G. Collman, Brendan J. Kelly, Kyle G. Rodino, Frederic D. Bushman, Roderick B. Gagne, Eman Anis

## Abstract

The SARS-CoV-2 pandemic likely began by viral spillover from animals to humans^1-3^; today multiple animal species are known to be susceptible to infection^4-8^. White-tailed deer, *Odocoileus virginianus* are infected in North America at substantial levels^9-11^, and genomic data suggests that a variant in deer may have spilled back to humans^12,13^. Here we characterize SARS-CoV-2 in deer from Pennsylvania (PA) sampled during fall and winter 2021. Of 123 nasal swab samples analyzed by RT-qPCR, 20 (16.3%) were positive for SARS-CoV-2. Seven whole-genome sequences were obtained, together with six more partial spike sequences. These annotated as alpha and delta variants, the first reported observations of these lineages in deer, documenting multiple new jumps from humans to deer. The alpha lineage persisted in deer after its displacement by delta in humans, and deer-derived alpha variants diverged significantly from those in humans, consistent with a distinctive evolutionary trajectory in deer.

## Main text

White-tailed deer were sampled throughout Pennsylvania from 10/2/2021 to 12/27/2021 with the goal of characterizing this potential new animal reservoir of SARS-CoV-2. Samples were tested for the presence of SARS-CoV-2 by qPCR, and the viral lineages present assessed using either viral whole genome sequencing after multiplex PCR, or, for samples with lower RNA amounts, sequencing of a single PCR amplicon encoding the spike receptor binding domain.

SARS-CoV-2 was detected by RT-qPCR in nasal swabs from 20 of 123 wild white-tailed deer sampled (16.3%; 95% CI: 10.4, 24.2) (described in Table S1 and S2). There was no significant difference in infection frequency associated with the cause of death (road kill 7/24, and hunter harvested 12/79; p-value = 0.14). There was no significant difference in infection rates between sexes (females 6/43, males 14/60; p-value = 0.46) and age groups (fawn 1/18, yearling 1/22, and adult 17/81; p-value = 0.093). No information was available on possible symptoms or disease for the animals sampled.

Virus-positive deer were identified in 10 of the 31 Pennsylvania counties surveyed (Figure 1); a Bayesian estimate of the infection rates is shown by the color scale. We estimate an average county has 9.9% positivity (95% CrI: 1.7-22%). Pike and Monroe counties were associated with higher apparent prevalence (Monroe: 7.5x higher odds (1.0-110x) of positivity; Pike: 9.3x higher odds (1.3-110x)). There was a significant difference in infection rates among regions (p-value = 0.0047), with the northeast region of PA showing a higher proportion of positive deer than the southeast region (Fisher’s exact test with FDR correction; p-value = 0.017).

**Figure 1.**
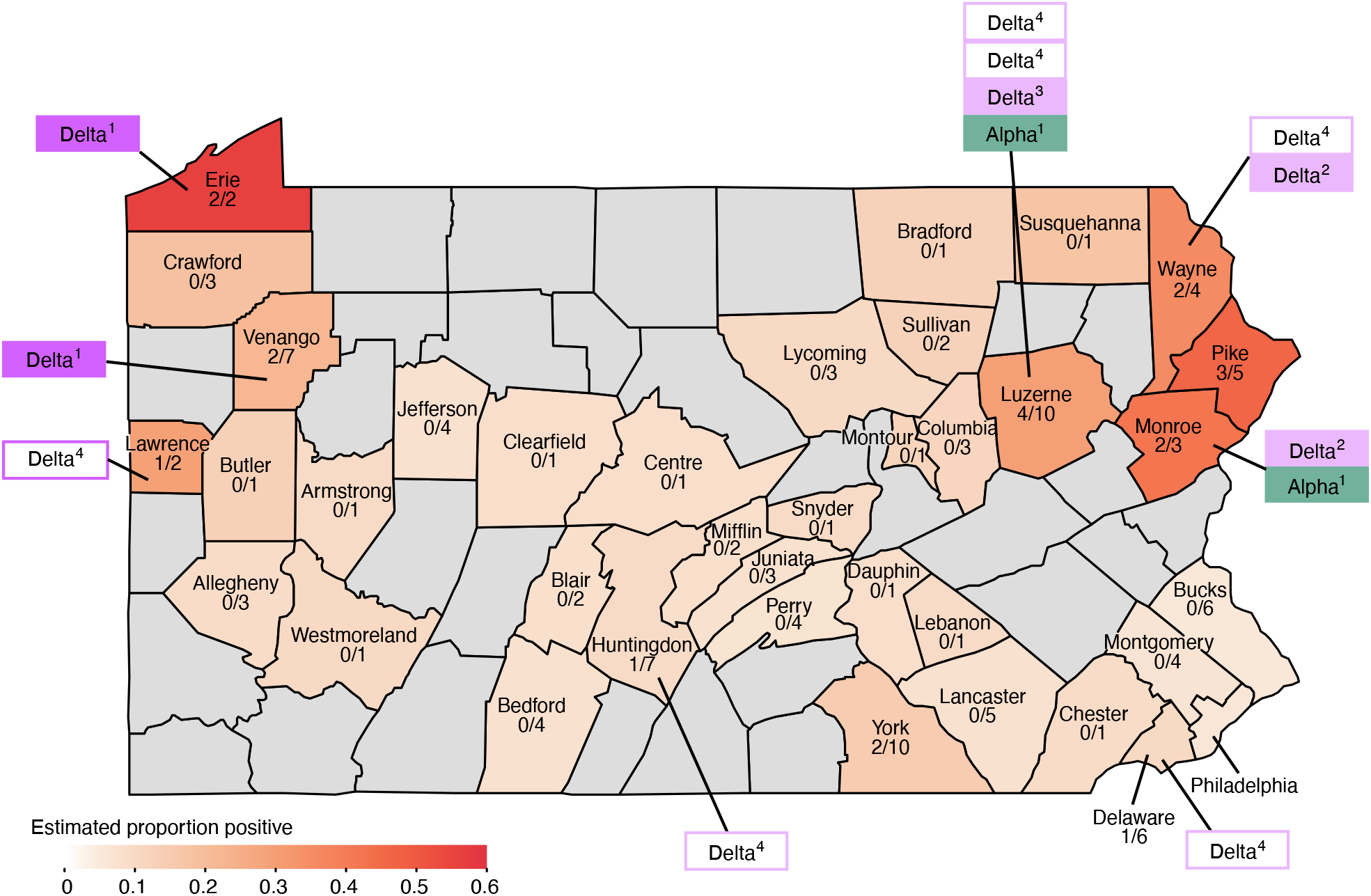
Map of Pennsylvania (PA), showing sampling sites and locations of SARS-CoV-2 positive deer. The counties comprising PA are outlined. The estimated proportion of positive samples is shown by the white-to-red color scale; counties that were not sampled are shown in grey. Total sample numbers and the number positive are written on each county sampled. The deer samples sequenced were assigned to variants as indicated by the rectangles outside the map; variant type is color coded (teal for alpha/B1.1.7, purple/pink for delta/AY.#). The open boxes indicate sequences were available for the spike only.

Viral whole-genome sequencing was attempted on eight samples with relatively high viral RNA concentrations (cycle threshold values from RT-qPCR less than 30, a cut value determined from analysis of human specimens), and high-quality genome sequences were recovered from seven (Table S2). To confirm sample provenance, the non-viral sequence reads were analyzed for the deer samples and for human samples that were sequenced in parallel (Table S6). All deer samples yielded reads mapping to the deer genome, and few or none mapping to the human genome. In contrast, reads from human samples overwhelmingly mapped to the human genome (Figure S1), confirming the host species origin of our samples as deer.

Samples with lower SARS-CoV-2 RNA concentrations were amplified by a single nested PCR amplification targeting the spike coding region as described^14^ and sequenced, yielding spike sequences from six additional samples. Spike sequences were then used for lineage assignment^15^ (Table S2).

Deer whole genome sequences showed divergences relative to the original Wuhan SARS-CoV-2 strain and previously reported human and deer genome sequences (Figure 2 A). A full list of deer substitutions is in Tables S3. The complete deer genome sequences reported here annotated as either alpha or delta variants^16^, the first reported identification of these lineages in wild white-tailed deer. Previously the alpha variant was shown to be able to infect white-tailed deer that were experimentally inoculated^11^. All the spike-only sequences harbored polymorphisms that place them in the delta lineage and were inconsistent with other common lineages.

**Figure 2.**
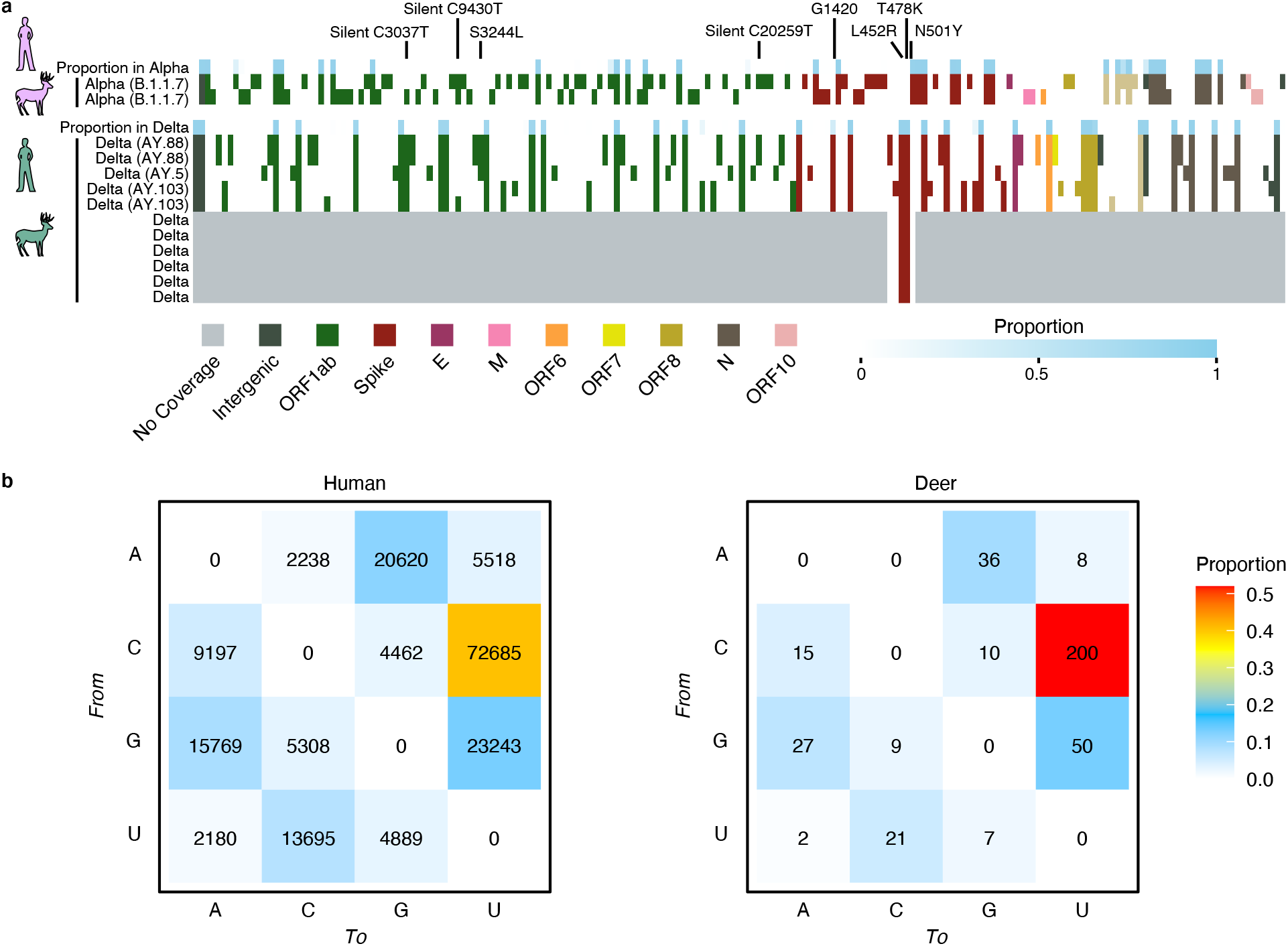
Base substitutions detected in sequences of deer from Pennsylvania. In A) the proportions in humans sampled in PA is shown in blue along the top for alpha (upper) and delta (lower). Substitutions relative to the Wuhan strain references are shown by the colored boxes. The proportions in humans are shown by the blue shading in each box. Genes sampled are shown by the color code indicated along the bottom. Grey indicates lack of sampling. The bottom six rows of the delta samples indicate the spike-only amplicon sequences. B) Types of base substitutions away from the Wuhan reference detected in humans (right) and deer (left). The proportions are shown by the color code at the left.

The types of base changes associated with substitutions was not uniform. Changes from C in the Wuhan reference to U were found to be the most frequent (Figure 2B). The frequency of C to U was even higher in deer than in humans (Pearson’s Chi-squared test p=0.031, followed by a post hoc analysis based on residuals yielding p-value = 0.0010 for C to U substitutions), consistent with a previous study^13^, and suggestive of host-specific mutation rates.

To identify any deer-specific sequence polymorphisms, the alpha and delta SARS-CoV-2 sequences from deer determined here were compared to viral genomes from humans and deer reported previously. All deer genome sequences available from GISAID (n=108) were downloaded (Table S4 and S5), corresponding to samples acquired from 9/28/2020 to 2/25/2021 in Iowa and Ohio and dominated by early pandemic lineages including B.1.2 and B.1.311^9,10^. Local human isolates were derived from our SARS-CoV-2 sequencing program monitoring the Delaware Valley, including Philadelphia, PA^17,18^. This revealed several substitutions that were highly enriched or invariant in deer isolates, but rare or absent in human isolates. These include three silent mutations in ORF1ab, C7303U, C9430U, and C20259U. Mutation C7303U was found in 86% of these PA deer and 29% of previously published deer, whereas it was found in 0.04% of Delaware Valley human subjects and 0.09% of global genomes reported by NextStrain. Mutation c9430t was found in 43% of PA deer and 56% of previously published deer, but 0.35% of Delaware Valley human subjects and 0.46% of global genomes. Mutation c20259t was found in 43% of PA deer and 19% of previously published deer, whereas it was absent in Delaware Valley human subjects and present in 0.12% of global genomes (data from 1/26/2022). The enrichment of these mutations suggests possible functional interaction with deer-specific factors, which could influence RNA synthesis, RNA folding, or protein binding.

We next analyzed the frequencies of synonymous and nonsynonymous substitutions in each genome to assess potential evolutionary pressures on SARS-CoV-2 in deer. The dN/dS ratios for the deer genomes did not diverge notably from matched viral variants in humans--the delta lineage in humans showed more evidence for diversifying selection than did alpha, and this was paralleled by sequences in deer. Strong diversifying selection was seen for the spike coding region in alpha and delta in both hosts, while neutral or purifying selection was more evident in most other coding regions (Supplementary Figure S2).

To assess relationships among deer and human-derived SARS-CoV-2 sequences, we used NextClade for sequence alignment and IQ-TREE to construct a maximum-likelihood phylogenetic tree for viral sequences from alpha (Figure 3A) and delta (Figure 3B) variants, in each case comparing PA deer to contemporaneous human-derived sequences from the same variant in the Delaware Valley region. We also compared the deer lineages observed longitudinally to all contemporaneous human lineages sampled in PA (Figure 3C, Table S7).

**Figure 3.**
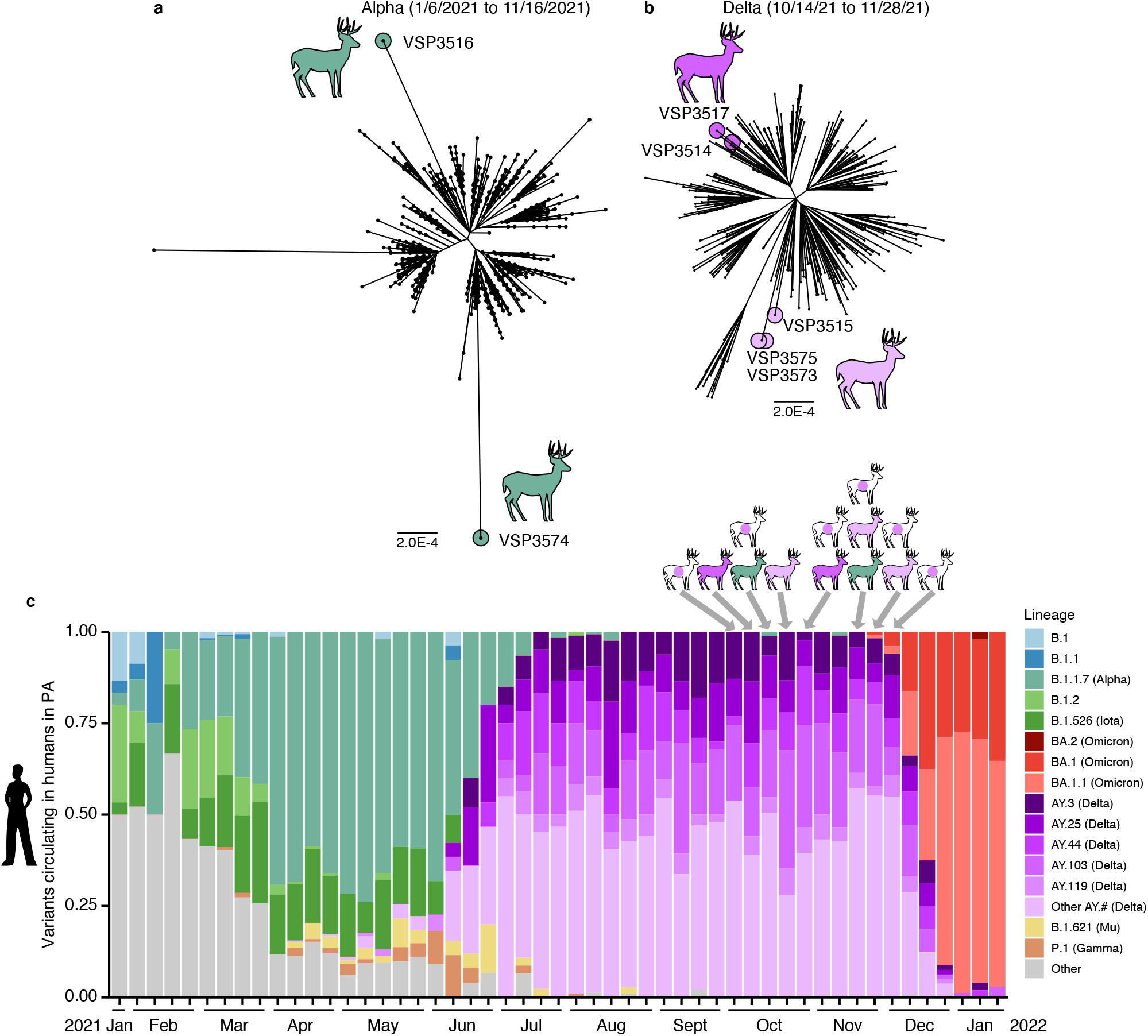
Analysis of SARS-CoV-2 whole genome sequences from white-tailed deer in PA and comparison to local human-derived sequences. A) Phylogeny of contemporaneous human alpha lineage sequences from PA, collected from 1/6/2021 to 11/16/2021, with deer sequences marked (1240 human and 2 deer sequences). B) Phylogeny of human delta lineage sequences from PA, collected from 10/14/2021 to 11/28/2021 (spanning the period of genomes obtained from deer), with deer sequences marked (440 human and 5 deer sequences). Time-resolved trees for each spillover are in Figure S3. C) Longitudinal comparison of deer variants and contemporary human variants. The bar plots show the progression of SARS-CoV-2 variants detected by genome sequencing in humans in eastern PA from 1/31/2021 to1/3/2022. The variants are color-coded according to the key to the right. The variants from white-tailed deer sequences are shown at the top of the figure, with the arrows showing the times of sampling. The deer isolates are color-coded on the deer icons as in the key; purple dots on deer icons indicate sequences identified as delta lineage by spike amplicon sequences but without whole genome sequences to further identify clade.

The two alpha sequences sampled from deer came from adjoining counties in northeastern PA. The isolates differed by 45 substitutions and were widely separated on the phylogenetic tree. A parsimonious explanation is that the two alpha lineages were introduced independently into deer and then diversified during transmission within deer. Only one out of 1,240 human genomes was as divergent or more divergent from the inferred alpha common ancestor as were the deer sequence, giving a probability of randomly drawing two such divergent lineages as (1/1240)^2^=6.5×10^−7^. Molecular clock analysis yielded data consistent with a model in which alpha jumped from humans to deer in April/May of 2021, during the alpha wave, and persisted in deer up until the sampling time six to seven months later in mid-November (Figure S3, Table S7 and S8).

For the delta variant (Figure 2B), five deer genomes annotated as delta, and all six spike sequences showed substitution patterns also consistent with delta (Table S3). Two deer genomes were assigned to the AY.103 clade, two genomes to AY.88, and one to AY.5. The delta lineages from deer were not as diverged from related human sequences as the alpha lineages, consistent with more recent introductions and less long-term circulation in deer. Molecular clock analysis suggested that the closely related AY.103 pair of genomes likely entered the deer population in mid-July 2021; the AY.5 lineage entered in mid-August 2021; and the AY.88 pair entered deer in late-September 2021. For the delta cases, the proposed jumps from deer to humans occurred during the height of the delta wave. The six samples sequenced as single spike amplicons also displayed mutations consistent with delta variants, and were collected at times and locations similar to the fully sequenced delta variant genomes from deer. The small size of these amplicons prevents assignment to clade and identification of transmission events but confirms the extensive presence of delta in deer.

Thus, these data support five independent jumps of SARS-CoV-2 from humans to deer—one each for the two alpha sequences, and three for the delta sequences (Supplementary Figure S3). Other scenarios are also possible, involving more independent jumps and convergent evolution in deer, or fewer jumps and divergent evolution along trajectories matching trajectories in humans.

This study has several limitations. The sample size is modest, and whole genome sequence acquisition was limited by viral RNA concentrations in samples. Our sample size limits the interpretation of epidemiological findings (e.g., differences in infection rates between sample types) but has less influence on the primary findings of this work (e.g., divergence of the alpha lineage in deer, and evidence for multiple spillovers). The background human data is incomplete, with only an estimated 1.7% percent of all human cases in PA subjected to viral whole genome sequencing. Each evolutionary analysis of sequences requires assumptions on the structures of background populations that are not fully investigated. In addition, we were not able to obtain serum samples from the deer studied and so could not investigate immune responses.

In summary, a survey of SARS-CoV-2 in 123 deer in Pennsylvania over the fall-winter of 2021 showed 16% of the deer sampled to be positive. Prior surveys carried out over the fall and winter of 2020 also showed high point prevalence of infection in Iowa (33%)^9^ and Ohio (36%)^10^, and later extensive infection at other locations ^13,19^. We report the first examples of the alpha and delta lineages in wild white-tail deer, likely derived from roughly five independent spillovers from humans to deer. Given that there are estimated to be 30 deer per square mile in PA, and over a million deer total, this suggests an enormous number of spillovers and infected deer in the state^20,21^. Mechanisms of infection and transmission are incompletely understood. Studies of experimental infections show efficient transmission between deer, potentially involving close interactions such as touching noses and grooming^22^. The mechanism of efficient transmission from humans to deer remains obscure. Our findings of alpha persistence in deer after replacement of alpha by delta in humans, and the divergence seen between our deer and human alpha genomes, are all consistent with long-term persistence and spread of the alpha variant in deer. As yet there is no evidence for spill-back of the deer lineages identified here into humans; ongoing efforts to characterize human and deer SARS-CoV-2 lineages are valuable to maintain surveillance for such events.

## Methods

### Collection of samples from white-tailed deer

Samples were collected from hunter-harvested deer and injured deer that were euthanized by state game wardens. Nasal swabs were taken within hours of death, placed in phosphate buffered saline (PBS) and stored in commercial refrigerators in field offices. Samples were shipped to the University of Pennsylvania within one week of collection and stored at −80°C until RNA extraction. Fisher’s Exact tests were performed using R Statistical software (v4.0.5)^23^ (R Core Development Team 2021) to test for differences in proportions of virus positive and negative deer by sex, age, and cause of death (i.e., hunter harvested, road-killed, or all other causes of death).

### RNA extraction and detection of SARS-CoV-2 RNA by RT-qPCR

Nucleic acid was extracted with a QIAamp viral RNA mini kit (Qiagen) according to the manufacturer’s instructions. The presence of SARS-CoV-2 RNA was assessed by a RT-qPCR targeting two regions of the viral nucleocapsid gene as described previously.^24^

### SARS-CoV-2 whole genome sequencing

Viral genomes were sequenced using the POLAR protocol^25^. The sample RNA (5μl) was mixed with Random Hexamers (0.5 μl of 50 μM, Thermo Fisher), dNTPs Mix (0.5μl of 10mM each, Thermo Fisher), and nuclease-free water (1 μl). This mixture was incubated (5 minutes at 65ºC). Subsequently, reverse transcription was performed adding 6.5 μl from the previous reaction to SuperScript III Reverse Transcriptase (0.5 μl, Thermo Fisher), 5X First-Strand Buffer (2 μl, Thermo Fisher), DTT (0.5 μl of 0.1M, Thermo Fisher), and RNaseOut (0.5 μl, Thermo Fisher). This mixture was incubated (50 minutes at 42ºC, then 10 minutes at 70ºC). For cDNA amplification, the previous mixture (2.5 μl) was added to Q5 Hot Start DNA Polymerase (0.25 μl, NEB), 5X Q5 Reaction Buffer (5 μl, NEB), dNTPs mix (0.5 μl of 10mM each, NEB), and pooled Artic-ncov2019 v4 primers (reaction set 1 used 4.0 μl of pooled primer set 1, reaction set 2 used 3.98 μl of pooled primer set 2, IDT) and water (volume reaction brought to 25 μl). The samples were brought to 98ºC for 30 seconds then cycled from 98ºC for 15 seconds to 65ºC for 5 minutes for 25 cycles, followed by 98ºC for 15 seconds and 65ºC for 5 minutes. The two reactions (one for each primer set) were pooled together. Tagmentation was completed with the Nextera XT Library Preparation Kit (Illumina). IDT for Illumina DNA/RNA UD Indexes were used for barcoding (Illumina). Quantification of DNA was performed using Quant-iT PicoGreen dsDNA quantitation assay kit (Invitrogen). The pooled libraries were quantified using the Qubit1X dsDNA HS Assay Kit (Invitrogen) and sequenced using the Illumina NextSeq platform.

### SARS-CoV-2 spike amplicon sequencing

Amplicon sequencing was carried out essentially as described^14^. Briefly samples with genomic viral load deemed too low for whole genome sequencing, or samples that failed whole genome sequencing, underwent nested PCR targeting the spike’s receptor binding domain sequence. Primers used are provided in Supplemental Table S9. PCR reaction 1 includes reverse transcription and initial DNA amplification using the Superscript IV One-Step RT-PCR System (Thermo Fisher Scientific,12594100). The 25 μl reaction consists of 5 μl of extracted viral RNA (extracted as described in the whole genome sequencing section), 0.25 μl of Superscript IV, 12.5 μl of 2X Platinum SuperFi RT-PCR mix, 1.25 μl of each PCR 1 primers (10 μM), 4.75 μl molecular grade water. The cycle conditions are as follows: 25ºC for 2 minutes, 50ºC for 20 minutes, 95ºC for 2 minutes, 25 cycles of amplification (95ºC for 2 minutes, 55ºC for 30 seconds, 70ºC for 1 minute). PCR reaction 2 is a nested PCR and addition of sequencing adapters for the amplicon product from reaction 1. The 25 μl mixture contains 5 μl of amplicon product from reaction 1, 12.5 μl 2x Q5 hot start master mix (NEB, M0494S), 0.5 μl of 10mM dNTPs (NEB, N0447S), 1.25 μl of each PCR 2 primers (10 μM), and 7.5 μl water. The conditions are as follows: 95ºC for 2 minutes, 20 cycles of amplification (95ºC for 15 seconds, 55ºC for 30 seconds, 72ºC for 1 minute). PCR reaction 3 adds the unique dual indexes and clustering adapters. The 50 μl reaction contains 25 μl of amplicon product from PCR reaction 2, 0.5 μl of Phusion polymerase (NEB, M0530L), 10 μl of 5x Phusion buffer, 1 μl of 10mM dNTPs (NEB, N0447S), 5 μl of IDT for Illumina DNA/RNA UD Indexes set A (Illumina, 20027213), and 8.5 μl of water. The reaction conditions are as follows: 98ºC for 3 minutes, 7 cycles of amplification (95ºC for 15 seconds, 50ºC for 30 seconds, 72ºC for 30 seconds), and 72ºC for 7 minutes. 10 μl of the barcoded PCR 3 mixture was pooled together, AMPure purified, and quantified by Qubit1X dsDNA and TAPE station before being sequenced on an Illumina MiSeq instrument using a 600 cycle v3 standard flow cell 290/10/10/290 protocol (Illumina, MS-102-3003). After sequencing, BWA v0.7.17-r1188 was used to generate bam files, Samtools v1.10 (using htslib 1.10.2-3) was used to filter reads for lengths between 280 and 300 base pairs, filter reads by quality using a PHRED threshold of 30, sort the reads, index the reads, and generate a pileup. Samples must meet a minimum mapping quality PHRED score of 30, have 95% 50-fold coverage for the 550bp amplicon target to pass additional filtering. Substitutions were called with a greater than 0.67 allele frequency for a given position.

### Sequence Data Processing

Sequence data were processed as previously described^17^. BWA aligner tool (v0.7.17) was used to align viral sequences to the Wuhan reference sequence (NC_045512.2)^26^. Samtools package (v1.10) was used to filter alignments^27^. Variants were called using Pangolin lineage software (3.1.17 with the PangoLEARN 2021-12-06 release)^16,28^. NVSL’s vSNP pipeline was also run on deer samples and results compared to those generated by our published pipeline; findings from both platforms deviated only for thresholds used to call low abundance substitutions. All variant calls remained the same between platforms.

### Host Sequence Analysis

The proportion of host sequences was inferred using a sampling of raw reads from all samples on any sequencing batch performed with deer specimen. 1,000 raw reads for each sample were blasted^29-31^ against a database constructed of a SARS-CoV-2 genome (NC_045512.2), human genome (GCF_000001405.39), feline genome (GCF_018350175.1), and white-tailed deer genome (GCF_002102435.1). Reads were tallied for each genome to which they were closely matched.

### Mutation Analysis

108 deer-derived SARS-CoV-2 genomes were downloaded from GISAID and compared with the 7 deer sequenced in this publication and the human dataset (Tables S4 to S6). The previously published genomes were downloaded on 12/29/2021. Mutations were called in reference to the Wuhan strain (NC_045512.2).

### Bayesian analysis of county proportions

To account for the variable sampling between counties and potential similarities between neighboring counties, we estimated the underlying proportion of deer testing positive within each county of the *m* counties using a Bayesian conditional autoregressive model. The number of positive tests, *y*_*i*_, out of *n*_*i*_ total tests within each county *i* was modeled as:

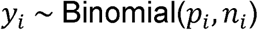

where *p*_*i*_ is the proportion of deer expected to test positive in that county and:

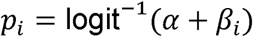

Here, *a* represents the average proportion positive for a county and the vector of differences from this average for each county, {*β*, is distributed as a multivariate normal:

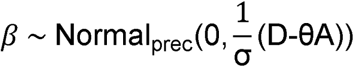

where *D* is a *m × m* matrix with 0s on the off diagonal and the diagonal element on each row *i, D*_*i,i*_, equal to the number of counties that are adjacent to county *i, A* is a *m × m* adjacency matrix with element *A*_*i,j*_ is 1 if county *i* neighbors county *J* and 0 otherwise and the diagonal set to 0 and Normal_prec_(*x*, *y*) is a multivariate normal distribution with means *x* and precision matrix *y*. For priors, θ was given a uniform prior between 0 and 1, σ ∼ Gamma(1,1) and α ∼ Normal(−2,10).

Posterior probability distributions were estimated using Markov chain Monte Carlo sampling using Stan v2.21.0 ^32^.

County adjacency data was obtained from the US Census bureau (https://www.census.gov/geographies/reference-files/2010/geo/county-adjacency.html).

### Analysis of dN/dS

Sequences were aligned using nextclade v1.14.0^33^. Coding sequences were extracted from the genomes ignoring insertions and treating ORF1a and ORF1b as separate entities. dN/dS was calculated from between individual gene sequences or the concatenation of all coding sequences for each genome against the ancestral Wuhan genome using the ape package ^34^ in R v4.1.3. Sequences with nonsense mutations and estimated dN/dS values less than 0 were discarded. Estimated dN/dS values >10 were censored to 10 and values <0.1 were censored to 0.1.

### Time-Scale Bayesian Maximum Clade Credibility Tree

A phylogenetic approach using a time-scale Bayesian maximum clade credibility (MCC) tree was used to estimate the introduction of SARS-CoV-2 into PA WTD. An NCBI BLASTn search was used to identify the nearest human-derived SARS-CoV-2 sequences for each deer, querying sequences world-wide and not just from the Delaware Valley. If a deer’s nearest hits included another deer, then both were analyzed in the same tree. Sequences without exact dates of collection were excluded. Sample sizes ranged from 50 to 79 nearest neighbor whole genome sequences. Each dataset was aligned using NextClade with Wuhan-Hu-1 as a reference (NC_045512.2). A Markov chain Monte Carlo (MCMC) method was used to generate time-scales Bayesian molecular clocks using BEAST v.1.10.4^35^. Several parameters were assessed to determine the tree best to model phylogenetic data using path sampling and stepping-stone sampling of marginal likelihood estimation (Supplemental Table S8). A substitution model described by Yang^36^, 1996 with an uncorrelated relaxed lognormal clock using Bayesian Skyline tree prior over 100 million iterations with subsampling every 1,000 iterations was deemed to be the optimal conditions over the parameters tested. The BEAGLE 3 library was used to improve computational performance^37^. Tracer v.1.7.1 was used to visually assess convergence. 100,000 iterations were discarded as burn-in. TreeAnnotator v.1.10.4 was used to summarize the MCC tree. FigTree v.1.4.4. was used to visualize the tree.

### Human subjects

Human sequences newly determined here were collected as follows. For most samples, the University of Pennsylvania Institutional Review Board (IRB) reviewed the research protocol and deemed the limited data elements extracted to be exempt from human subject research per 45 CFR 46.104, category 4 (IRB #848605). For hospitalized subjects, following informed consent (IRB protocol #823392), patients were sampled by collection of saliva, oropharyngeal and/or nasopharyngeal swabs, or endotracheal aspirates if intubated, as previously described^17^. Further samples were collected from asymptomatic subjects detected in a screening program at the Perelman School of Medicine at the University of Pennsylvania and symptomatic subjects tested throughout the PennMedicine clinical network under IRB protocols #843565 and #848608. Human samples were sequenced as described for deer samples.

### Phylogenetic Analysis

Deer and human sequences are available at GenBank (Table S2 and Table S6). NextClade was used to align sequences to the Wuhan reference^33^. IQ-TREE (v1.6.12) was used to infer a tree using maximum-likelihood methods using 1,000 bootstrap replicates^38^. Visualization of the inferred tree was performed using FigTree (v.1.4.4)^39^. Only a small fraction of human cases from Pennsylvania have been analyzed by viral whole genome sequencing (estimated at about 1.7% comparing GISAID upload for PA to state data on infection rates); thus, our background human data set for comparison to the deer samples is quite incomplete.

### Materials

Key materials and resources are compiled in Table S9.

## Data Availability

Sequence accession numbers for deer-derived SARS-CoV-2 genomes can be found in Table S3 (OM570187-OM570193). Accession numbers for human SARS-CoV-2 genomes can be found in Table S7. Computer code is available at https://doi.org/10.5281/zenodo.4046252.

## Data availability

Sequence accession numbers for deer-derived SARS-CoV-2 genomes can be found in Table S2 (OM570187-OM570193 & ON350842-ON350847). Accession numbers for human SARS-CoV-2 genomes can be found in Tables S6 and S7. Computer code is available at https://doi.org/10.5281/zenodo.4046252.

## Acknowledgements

We are grateful to hunters and wildlife personnel who provided specimens, and to Laurie Zimmerman for artwork and help with manuscript preparation. We acknowledge help from staff of the Philadelphia Department of Public Health. This work was supported in part by the Penn University Research Foundation. SG is supported by the Robert J. Kleberg, Jr. and Helen C. Kleberg Foundation. Funding was provided by a contract award from the Centers for Disease Control and Prevention (CDC BAA 200-2021-10986 and 75D30121C11102/000HCVL1-2021-55232), philanthropic donations to the Penn Center for Research on Coronaviruses and Other Emerging Pathogens, and in part by NIH grant R61/33-HL137063 and AI140442 -supplement for SARS-CoV-2. BJK is supported by NIH K23 AI 121485. Additional assistance was provided by the Penn Center for AIDS Research (P30-AI045008). This project has been funded in part with Federal funds from the National Institute of Allergy and Infectious Diseases, National Institutes of Health, Department of Health and Human Services, under Contract No. 75N93021C00015. HZ’s stipend to conduct this work was provided by a private donation by Tracy Holmes.

## Contributions

Conceived and designed the experiments. A. D. M., J. C. E., R. G. C., B. J. K., K. G. R., F. D. B., R. B. G., E. A.

Performed the experiments A. D. M., H. A., S. R., H. A., B. G., E. A.

Analyzed the data A. D. M, S. S.-M., J. E., F. D. B., B. G., E. A., S. G.

Contributed materials/analysis tools J. C. E., H. Z., S. S. G., C. C., K. S., R. G. C., B. J. K., K. G. R., F. D. B., R. B. G., E. A.

Wrote the paper A. D. M., S. S.-M., J. C. E., H. Z., S. S. G., C. C., K. S., R. G.

C., B. J. K., K. G. R., F. D. B., R. B. G., E. A.

## Supplementary Material

Supplementary Table 1. Table S1. SARS-CoV-2 prevalence estimates stratified by sex, age, sampling region, and cause of death.

Supplementary Table 2. Results of qPCR and sequencing analysis. Supplementary Table 3. Deer SARS-CoV-2 substitutions from sequences in this study.

Supplementary Table 4. Single nucleotide polymorphisms from global whole genome sequencing of SARS-CoV-2 in deer.

Supplementary Table 5. GISAID Acknowledgements for deer sequences.

Supplementary Table 6. Human sequences used for Figure 3C.

Supplementary Table 7. Deer and human sequences used for the time-scale Bayesian maximum credibility tree.

Supplementary Table 8. Parameter tuning for time-scale Bayesian maximum clade credibility tree.

Supplementary Table 9. Key materials and resources.

Supplementary Figure 1. Checking sample tracking by analysis of nonviral sequences. Sequence reads from each SARS-CoV-2 sample were aligned to the white-tailed deer (GCF_002102435.1) and human (GCF_000001405.39) genomes, and nonviral reads enumerated. The numbers of deer and human reads are shown for each deer sample, denoted by the laboratory accession number VSP###. Results for 140 human samples, sequenced in the same sample batches, are shown to the far left. The proportion of reads aligning to the deer genome are shown in green, the average fraction aligning to the human genome is shown in orange.

Supplementary Figure 2. Results of quantification of dN/dS ratios. Values are shown in each panel from right to left dN/dS in deer, in human alpha, human delta, and all other lineages. Lineages in deer are color coded as in the human samples. The dN/dS values were calculated comparing each sequence to the reference Wuhan strain and are shown for the concatenation of all open reading frames (upper left), ORF1a (upper right), ORF 1b (lower left), and Spike (lower right).

Supplementary Figure 3. Time resolved phylogenies, comparing deer and human lineages. Deer isolates are compared to human isolates from an overlapping geographic region. Trees are shown for individual lineages when relatively divergent from other deer isolates, or for pairs of deer isolates when similar in sequence.

