## Supplementary figures and images for "Multiple Introductions of SARS-CoV-2 Alpha and Delta Variants into White-Tailed Deer in Pennsylvania"

### Suppl Figure 1

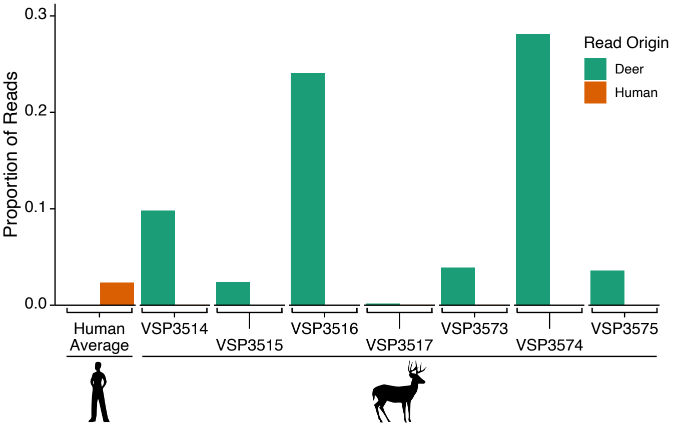

### Suppl Figure 2

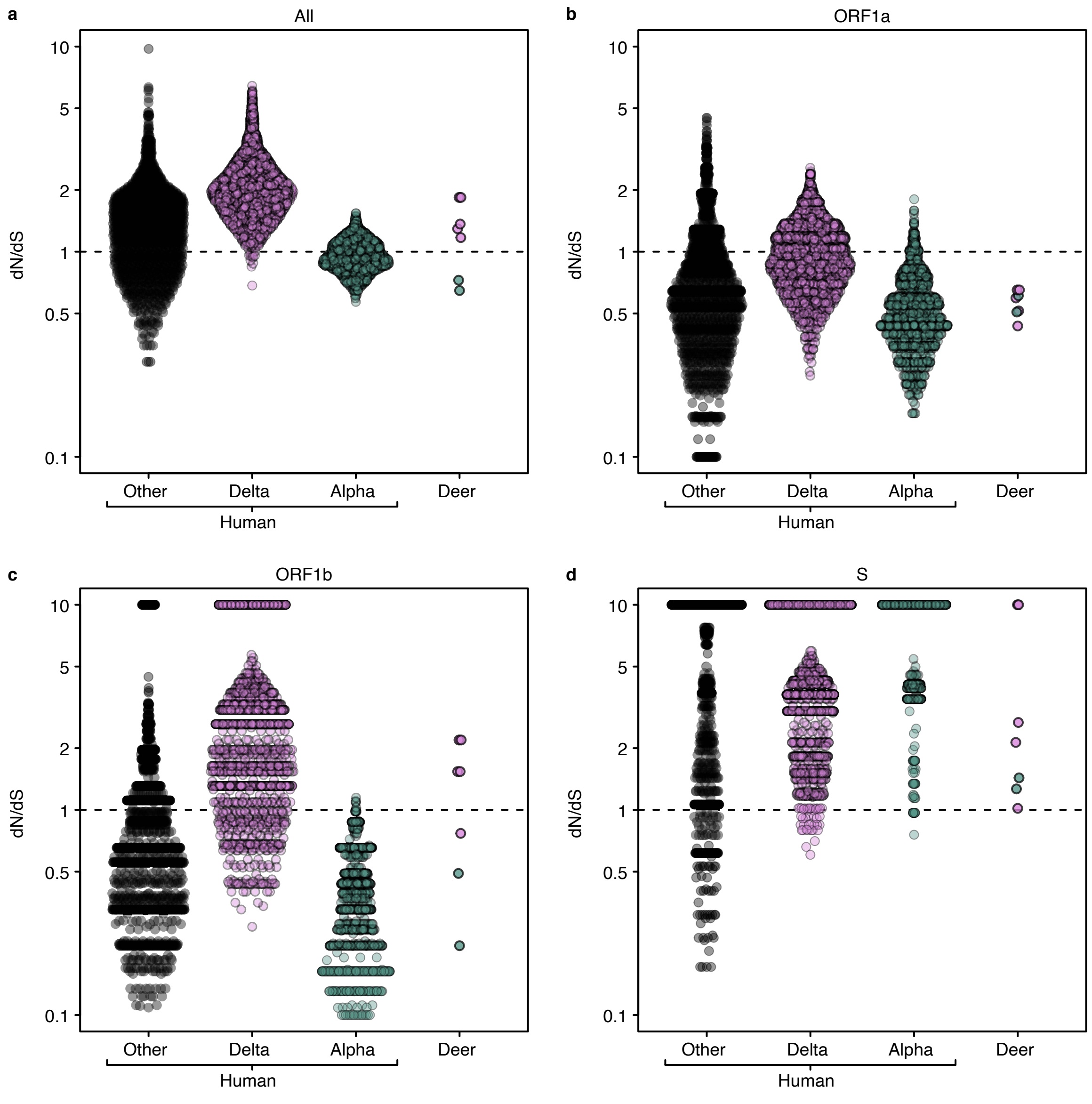

### Suppl Figure 3

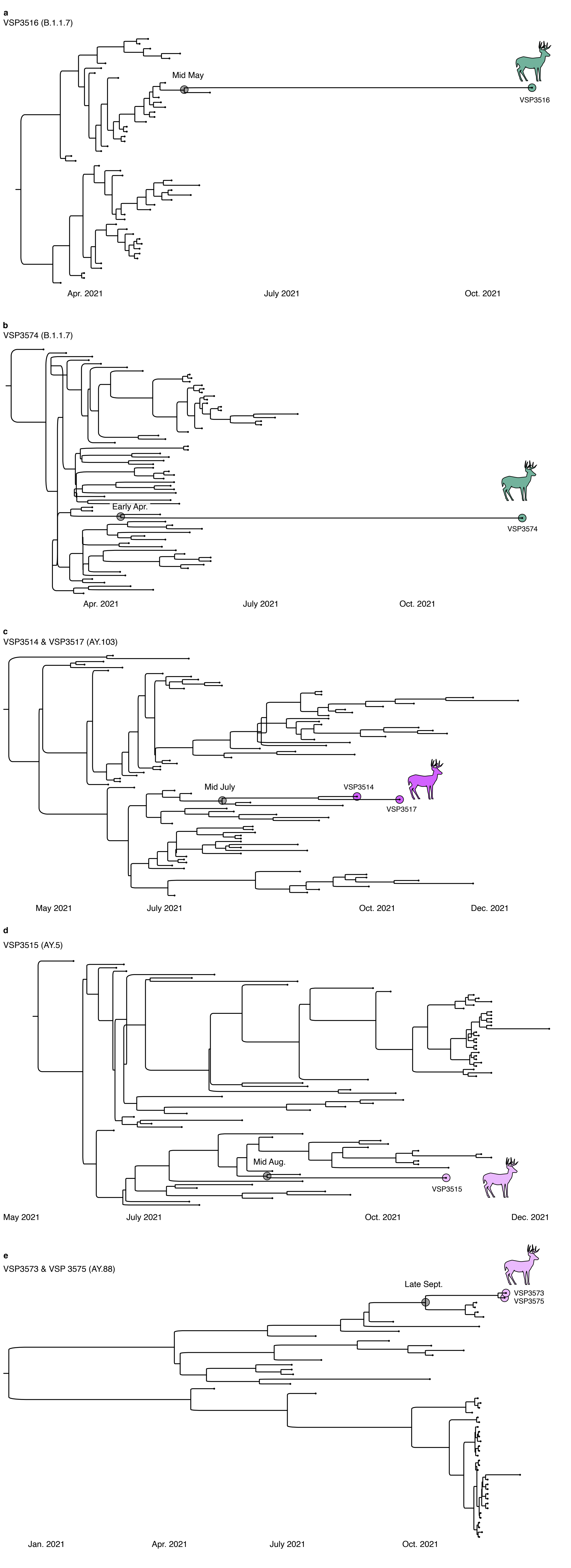
